# Mosaic chromosomal alterations in blood are associated with an increased risk of Alzheimer’s disease

**DOI:** 10.1101/2025.05.29.25328544

**Authors:** Tatsuhiko Naito, Kosei Hirata, Beomjin Jang, Chirag M. Lakhani, Alice Buonfiglioli, Wan-Ping Lee, Otto Valladares, Li-San Wang, Yukinori Okada, Hong-Hee Won, Eirini P. Papapetrou, Samuele G. Marro, David A. Knowles, Towfique Raj

**Author notes:** Corresponding author: These authors jointly directed this study, Tatsuhiko Naito, MD, Ph.D. Departments of Neuroscience, and Genetics and Genomic Science, Icahn School of Medicine at Mount Sinai, New York, NY, Towfique Raj, Ph.D., Departments of Neuroscience, and Genetics and Genomic Science, Icahn School of Medicine at Mount Sinai, New York, NY.

## Abstract

Mosaic chromosomal alterations (mCAs) in blood, a form of clonal hematopoiesis, have been linked to various diseases, but their role in Alzheimer’s disease (AD) remains unclear. We analyzed blood whole-genome sequencing (WGS) data from 24,049 individuals in the Alzheimer’s Disease Sequencing Project and found that autosomal mCAs were significantly associated with increased AD risk (odds ratio = 1.27; *P* = 1.3 × 10^−5^). This association varied by ancestry, mCA subtype, APOE ε4 allele status, and chromosomal location. Using matched blood WGS and brain single-nucleus RNA-seq data, we identified microglia-annotated cells in the brain carrying the same mCAs found in blood. These findings suggest that blood mCAs may contribute to AD pathogenesis, potentially through infiltration into the brain and influencing local immune response.

## Main

Clonal hematopoiesis (CH) is a form of age-related somatic mosaicism in the hematopoietic system characterized by the presence of clonally expanded hematopoietic stem or progenitor cells and their progeny harboring somatic mutations in driver genes associated with myeloid malignancies.^1^ CH is categorized into several primary types, including autosomal mosaic chromosomal alterations (mCAs), loss of sex X and Y chromosomes (LOX and LOY), and clonal hematopoiesis of indeterminate potential (CHIP).^2^ As these are founding mutations for hematological neoplasms, CH is associated with a higher risk of developing cancers.^3^ Additionally, CH has been linked to range of other diseases, such as cardiovascular and respiratory diseases.^4, 5^ For neurodegenerative diseases, LOY was previously reported to be significantly associated with the risk of Alzheimer’s disease (AD).^6^ More recently, a protective effect of CHIP against AD was reported by analyzing multiple cohorts.^7^ However, the burden of CHIP and mCAs in brain cells, such as microglia, has been reported to contribute to an increased risk of AD.^8^ These distinct effects of different CH classes motivate deeper investigation into the relationship between CH and AD. In particular, little is known about the involvement of autosomal mCAs in blood in the risk of AD. Recently, technological advances have enabled the detection of not only CHIP,^9^ but also mCAs from whole-genome sequencing (WGS) with a higher sensitivity than genotyping arrays.^10^ Here, we explore the association of autosomal mCAs in blood, including copy number gains and losses, and copy number neutral loss of heterozygosity (CN-LOH), with AD using WGS of a large multi-ancestry cohort of the Alzheimer’s Disease Sequencing Project (ADSP) consortium.^11, 12^

After applying our inclusion criteria and quality control measures (see Methods), we analyzed 8,288 AD cases and 15,761 controls with mean ages of 72.0±10.3 and 73.0±9.1 years, respectively (**Supplementary Table 1**). We found that 8.1% of individuals had at least one mCA of any type, with chromosomal distributions consistent with those reported in previous studies (**Supplementary Fig. 1**). The prevalence of mCAs increased with age (**Fig. 1a**). The presence of any mCA was significantly associated with AD in logistic regression analysis (odds ratio [OR] = 1.26, 95% confidence interval [CI] = 1.13−1.40, *P* = 2.0 × 10^−5^; **Fig. 1b** and **Supplementary Table 2**). The effect of mCAs on AD appeared to vary by ancestry group, with the strongest association observed in African American (AFR) individuals (OR = 1.45, 95% CI = 1.18−1.78, *P* = 4.0 × 10^−4^; **Fig. 1b** and **Supplementary Table 2**), possibly reflecting differences in mCA distribution as well as heterogeneity in AD phenotypes and etiology. We next examined the association of each mCA type compared to individuals without any mCA. CN-LOH showed the strongest association (OR = 1.40, 95% CI = 1.21−1.63, *P* = 8.8 × 10^−6^; **Fig. 1b** and **Supplementary Table 2**), with nominally significant association in all separated ancestries (*P* < 0.05).

**Figure 1.**
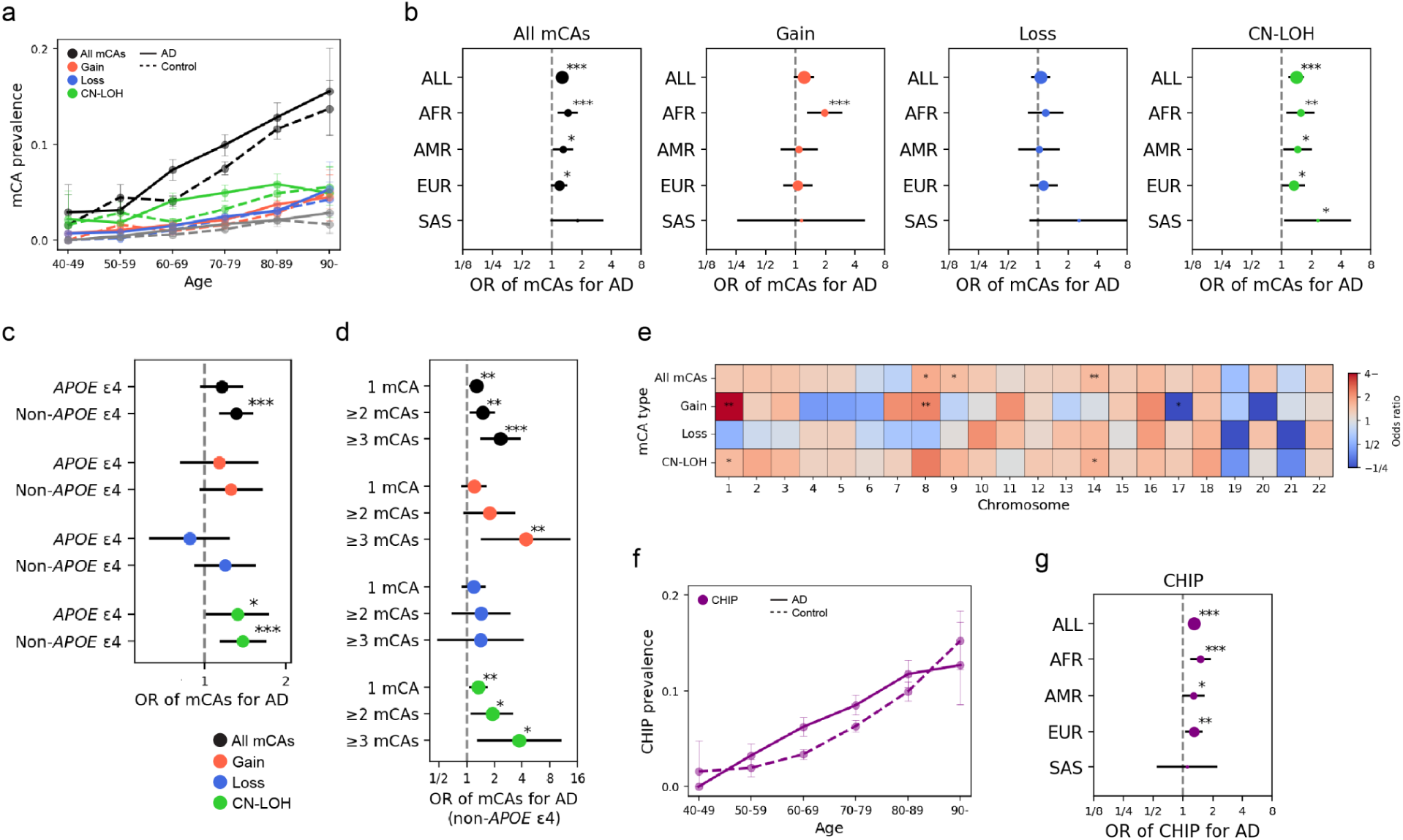
The association between autosomal mCAs and AD. **a.** The prevalence of mCAs by age. The solid and dashed lines represent AD cases and controls, respectively. Colors indicate mCA types. **b.** Forest plots showing the odds ratio (ORs) of mCAs for AD in each ancestry group, with results presented by mCA type. **c.** Forest plot showing the ORs of mCAs for AD in individuals with and without the *APOE* ε4 allele. **d.** Forest plot showing the ORs for the presence of 1, 2, or ≥3 mCAs for AD in individuals without the APOE ε4 allele. **e.** Heatmap showing the ORs of mCAs on each chromosome for AD. **f.** The prevalence of CHIP by age. The solid and dashed lines represent AD cases and controls, respectively. **g.** Forest plot showing the ORs of CHIP for AD in each ancestry group. In all plots, the error bars represent 95% confidence intervals. *, **, and *** represent *P*-value in association test < 0.05, 0.01, and 0.001, respectively. ALL, all ancestries; AFR, African American; AMR, Admixed American; EUR, European; SAS, South Asian; AD, Alzheimer’s disease; OR, odds ratio.

We investigated whether the association between mCAs and AD varied by *APOE* ε4 genotype, a major genetic risk factor for AD that is associated with different phenotypic and pathological features.^13^ The mCA-associated risk tested in all ancestries combined appeared to be stronger among *APOE* ε4 non-carriers (OR = 1.16 vs 1.31 for the presence of any mCA types between APOE4 carriers and non-carriers in all ancestries; **Fig. 1c** and **Supplementary Table 3**). However, when stratified by ancestry, the tendency varied, with stronger associations observed among *APOE* ε4 non-carriers in Europeans (EUR) (OR = 1.00 vs 1.32, *P*_int_ = 0.033), whereas in South Asians (SAS), stronger associations were seen among *APOE* ε4 carriers (OR = 6.73 vs 1.16, *P*_int_ = 0.021; **Supplementary Fig. 2** and **Supplementary Table 3**). We further examined the impact of multiple mCAs and found that the associations became stronger with an increasing number of mCAs, particularly among *APOE* ε4 non-carriers (**Fig. 1d** and **Supplementary Table 4**), suggesting a mutational burden effect of mCAs on the risk of AD.^10^ To formally test this trend, we included the count of mCAs, instead of a binary indicator of mCA presence, in the model, restricted to individuals carrying mCAs. The trend was significant in *APOE* ε4 non-carriers (*P* = 0.029). Finally, we analyzed mCAs separately by chromosomal location to determine whether specific genomic regions were more strongly associated with AD. Our results suggested regional heterogeneity, with the gain on chromosomes 1 and 8 showing the most significant association (OR = 5.88, 95% CI = 1.68−20.53, *P* = 0.0055 for chromosome 1 and OR = 2.45, 95% CI = 1.40−4.42, *P* = 0.0052 for chromosome 8; **Fig. 1e** and **Supplementary Table 5**).

Given the previously reported co-occurrence of mCAs and CHIP,^10^ we investigated their combined effect on AD risk by detecting individuals harboring CHIP from the same blood WGS data. We found that 6.9% of individuals carried at least one CHIP event, and the prevalence of CHIP increased with age (**Fig. 1f**). In our data, CHIP was significantly associated with an increased AD risk (OR = 1.30, 95% CI = 1.16−1.46, *P* = 4.2 × 10^−6^; **Fig. 1g** and **Supplementary Table 1**). When both mCAs and CHIP were included in the same model, each remained independently associated with AD (*P* = 5.1 × 10^−5^ and 1.1 × 10^−5^, respectively; **Supplementary Fig. 3** and **Supplementary Table 6**), with no significant interaction effect detected (*P* = 0.88). This independent association of mCA was observable even in previously reported region-level co-occurrence patterns, such as the effect of gain on chromosome 8 conditioned on CHIP of *DNMT3A* (*P* = 0.0061).^10^

The previous study of CHIP in AD reported the potential presence of microglia-annotated cells having the same CHIP mutations as identified in blood, suggesting bone marrow-derived monocytes with CH may infiltrate into the brain and contribute to AD development.^7, 14^ Using matched blood WGS and brain single-nucleus RNA-seq (snRNA-seq) data, we investigated the presence of microglia-annotated cells with the same mCA as in blood for the Religious Orders Study/Memory and Aging Project (ROS/MAP),^15^ (**Fig. 2a**). We utilized two partially overlapping snRNA-seq datasets from the ROS/MAP cohort, designating one as the primary dataset^16^ and the other as a replication set.^17^ In the primary dataset,^16^ we identified 11 individuals with mCAs in blood among 145 unique individuals analyzed. We then applied Numbat, a copy number variant (CNV) detection tool for single-cell sequencing data that estimates the probability of each cell harboring a CNV utilizing pre-specified CNV locations as prior information.^18^ To support stringent detection of mCA-positive cells in snRNA-seq data, despite inherent technical noise and challenges of CNV calling, we tested whether cells carrying the same CNVs as those observed in blood were enriched among microglia-annotated cells (vs other annotated CNS cell types), under the assumption that only microglia-annotated cells originate from blood among the annotated cell populations. We identified four individuals with a marked enrichment of cells harboring the same mCA as detected in blood specifically within microglia-annotated cells, with a positive correlation in mCA cell fractions between blood and microglia-annotated cells (Spearman’s ρ = 0.90, *P* = 0.037; **Fig. 2a, b** and **Supplementary Table 7**). Within previously defined microglial states,^19, 20^ including MG1 (surveilling genes), MG2 (reactive genes), MG3 (redox-related genes), and MG4 (lipid-processing genes), microglia-annotated cells harboring mCAs were significantly enriched in MG3 (OR = 2.26, 95% CI = 1.27−4.01, *P*_adj_ = 0.011) and depleted in MG4 (OR = 0.44, 95% CI = 0.27−0.71, *P*_adj_ = 0.0036; **Fig. 2c, d**). In the replication dataset, we identified significant enrichment of cells harboring the same mCA as detected in blood within microglia-annotated cells in four individuals among 11 individuals with blood mCAs, with the mCA of one of the shared individuals replicated, despite a less clear correlation in mCA cell fractions (Spearman’s ρ = 0.20, *P* = 0.8; **Supplementary Fig. 4a** and **Supplementary Table 7**). The enrichment pattern of microglial states was replicated with the enrichment in MG3 (OR = 3.43, 95% CI = 2.02−5.84, *P*_adj_ = 1.0 × 10^−5^) and with the complete depletion in MG4 (**Supplementary Fig. 4b, c**). These findings suggest that these microglia-annotated cells harboring mCAs are functionally impaired, with mCAs potentially predisposing them to disrupted phagocytic capacity in early states, which may contribute to AD risk,^21, 22^ or progression.

**Figure 2.**
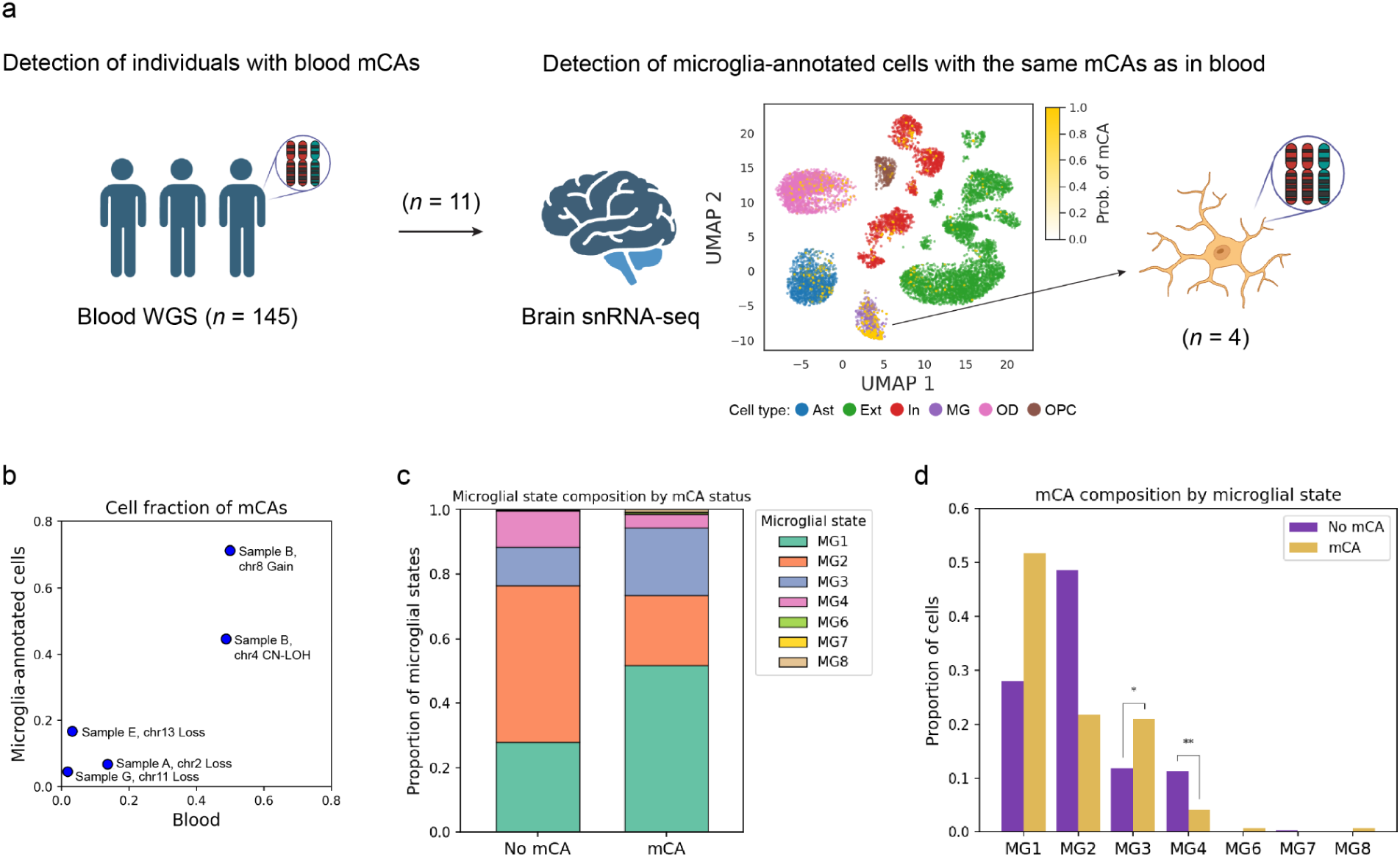
Detection of the same mCA as identified in blood from brain snRNA-seq data. **a.** Using individuals with paired blood WGS and brain snRNA-seq data, we first identified those carrying blood mCAs, and then detected cells harboring the same mCAs, which were markedly enriched within the microglia-annotated population. The central uniform manifold approximation and projections (UMAP) plot shows the probability of harboring mCAs overlaid on cell type annotations in samples where mCAs were significantly enriched in microglia-annotated cells. **b.** Scatter plot showing the cell fractions of mCAs in blood and microglia-annotated cells. **c.** Bar plot showing the proportion of microglial states by mCA status. **d.** Bar plot showing the proportion of cell with and without mCA across different microglial states. Some illustrations were made with the aid of BioRender. Ast, astrocytes; Ext, excitatory neurons; In, inhibitory neurons; MG, microglia; OD, oligodendroglia; OPC, oligodendrocyte progenitor cells.

Here, by analysing WGS from a large cohort, we provided findings supporting the risk association of autosomal mCAs in blood with AD. In particular, our observation that microglia-annotated cells harbor the same mCA as identified in blood samples raises the possibility that monocytes with mCAs might enter the brain,^7^ in line with the previous study of CHIP mutations.^7^ Furthermore, if monocytes with mCA could enter the brain and acquire features of microglia, our results align with a previous study demonstrating that mutational burden, including mCAs, in microglia contributes to an increased risk of AD.^8^ It remains unknown how microglia with mCAs contribute to risk; however, bone-marrow-derived monocytes, which are generally thought to be protective in AD,^14^ may be functionally impaired.

Our study has several limitations and suggests directions for future research. First, the analysis was based on a single consortium-based dataset, although our ancestry-stratified design provided reciprocal replication of the associations. Second, the study is cross-sectional and observational, which cannot exclude the possibility of reverse causation, although there is no clear rationale to suggest that AD causes mCAs. Longitudinal follow-up studies will be necessary to more rigorously support a causal relationship between mCAs and AD. Finally, in the exploration of microglia-annotated cells harboring mCAs, contamination by circulating blood cells cannot be entirely ruled out. Functional validation will be important to clarify the biological relevance of these findings.

## Code availability

The analysis steps, functions, and parameters used are described in the Methods section. Custom scripts used for data analysis are available upon reasonable request.

## Data Availability

This paper uses the ADSP Release 4 WGS data and AD phenotype data.

## Acknowledgments

We thank the patients and families who donated material for these studies. Research reported in this paper was supported by the Alzheimer’s Disease Sequencing Project of the National Institutes of Health under award numbers U01 AG068880. T.R. and T.N. were supported by National Institutes of Health (NIH) grants, including NIA R56-AG088669, U01-AG058635, NIA R21-AG063130, NIA R01-AG054005, NIA U01-AG068880, NIA RF1-AG065926, NIA R56-AG055824, NIA P30-AG066514, NINDS U54-NS123743, and NINDS R01-NS116006. The content is solely the responsibility of the authors and does not necessarily represent the official views of the National Institutes of Health. T.N. was supported by the BrightFocus Foundation Postdoctoral Fellowship and the Japan Society for the Promotion of Science Overseas Research Fellowships.

This work was supported in part through the computational resources and staff expertise provided by Scientific Computing at the Icahn School of Medicine at Mount Sinai and supported by the Clinical and Translational Science Awards (CTSA) grant UL1TR004419 from the National Center for Advancing Translational Sciences. Research reported in this paper was supported by the Office of Research Infrastructure of the National Institutes of Health under award number S10OD026880 and S10OD030463.

## Author contributions

T.N. and T.R. conceived and designed the study and wrote the manuscript. T.N. analyzed the data and performed the statistical analyses with assistance from H.K. and B.J. C.M.L. and D.A.K. managed the ADSP data. W.-P.L, O.V., and L.-S.W. provided the ADSP data. Y.O., H.-H.W., A.B., E.P.P, S.G.M. and D.A.K. provided critical review and suggestions for the study and manuscript. All authors have read and approved the final manuscript. T.R. supervised the project.

## Ethics declarations Competing interests

The authors declare no conflicts of interest for this study. T.R. served as a scientific advisor for Merck and serves as a consultant for Curie.Bio.

## Methods

### ADSP consortium WGS data and clinical information

We analyzed the release R4 of WGS data from the Alzheimer’s Disease Sequencing Project (ADSP), consisting of approximately 36,000 samples from three main cohorts, which include individuals from different ancestries, such as African American (AFR), admixed American (AMR), East Asian (EAS), European (EUR), and South Asian (SAS).^11, 12^ The mean read depth across samples was 40.4x with 99% of samples having a coverage >30x.^11, 12^ Following the quality control (QC) performed by the consortium, we conducted additional processing of the WGS data. First, we combined phenotype information across multiple cohorts and removed genetically identical duplicates (IBD π^ > 0.95) and technical replicate samples, selecting those with the highest call rates. We prioritized individuals in family studies over those in case-control studies when they overlapped. Related individuals were removed using KING,^23^ keeping AD cases where possible, to prevent potential bias due to genetic predisposition to CH.^9^ We included only blood samples because the purpose of this study was to explore blood mCAs, while the dataset also contains other tissue data. We focused only on individuals aged over 40. We removed samples with missing data used in this study. We excluded EAS individuals due to the small sample size (*n* = 94). Consequently, we obtained 8,296 AD cases and 15,795 controls. The demographic characteristics of the individuals used in this study are summarized in **Supplementary Table 1**.

### Detection of autosomal mCAs

We detected autosomal mCAs from WGS using MoChA (v1.20) based on previously described pipelines.^10^ We used the WGS files generated and QC’d by the consortium. We phased the genotypes using SHAPEIT (5.1.0).^14^ We removed heterozygous markers with a minor allele frequency < 1%, those where the read depth of either allele was less than five, and markers within germline copy number variants. The MoChA caller was run with the extra option ‘–min-dist 1000 –LRR-weight 0.0 –bdev–LRR–BAF 6.0’ to disable the LRR + BAF model. The resulting mCA calls were filtered by excluding (1) those that span less than 2,000 informative markers, that is heterozygous sites; (2) those with logarithm of the odds score less than 5; (3) those with estimated relative coverage higher than 2.9; and (4) those with BAF deviation larger than 0.16 and relative coverage higher than 2.5. Consequently, we identified individuals having mCA in different categories, including “Gain”, “Loss”, “CN-LOH”, and “Undetermined”, in chromosomal segments. We confirmed that the chromosomal distributions of mCA in our data were consistent with those in the original study that developed the pipelines (**Supplementary Fig. 1**).^10^

### Detection of CHIP

We detected CHIP from WGS CRAM files using Mutect2 provided by GATK (v4.3.0.0) with default settings in Tumor-Normal mode based on a previously described pipeline.^7^ Raw variants called by Mutect2 were filtered using FilterMutectCalls, incorporating the estimated prior probability of a read orientation artifact generated by LearnReadOrientationModel. Putative variants flagged as “PASS” by FilterMutectCalls or flagged as ‘germline’ if present at least two times with the “PASS” flag in other samples were selected for further analysis. Additionally, Mutect2 excludes common error variants observed in specific cohorts using a reference Panel of Normals, which we compiled from individuals under 40. The output from the Mutect2 pipeline was then annotated using Ensembl Variant Effect Predictor (v112)^24^ for detecting known CHIP variants in 73 genes from a curated list used in a previous study.^7^

### Association analysis between CH and AD

We performed a logistic regression analysis on the case-control status for AD, modeling CH statuses as variables. Age, age^2^, sex, age × sex, main cohort, sub-cohort, sequencing platforms, sequencing centers, the top ten principal components from genome-wide germline genotypes, and the combinations of *APOE* genotypes were included in the model as covariates. For the ancestry-combined analysis, we further included the predicted ancestry category to robustly adjust for ancestry bias. The statistical analyses were conducted using Statsmodels (v0.14.0), a Python library.

### Religious Orders Study/Memory and Aging Project (ROS/MAP) snRNA-seq data

We used brain snRNA-seq from the ROS/MAP^15^ cohorts to detect mCAs in microglia-annotated cells and perform subsequent analyses. Among the two studies with partial overlap in individuals, Fujita et al.^16^ and Mathys et al.^17^, we prioritized Fujita et al. due to its higher average total UMI count, while Mathys et al. was used as a replication dataset.

The details of snRNA-seq processing and quality control were described previously.^20^ Briefly, we performed the analysis using SCANPY (v1.9.8).^25^ Outlier cells (3 × IQR for gene count, total UMI, or mitochondrial content) were excluded, and doublets were removed using Scrublet^26^ (v0.2.3). Batch effects were corrected with Harmony^27^ using individual and batch covariates. Major cell types were annotated based on original studies.^16, 17^ In this study, we targeted astrocytes (Ast), excitatory neurons (Ext), inhibitory neurons (In), microglia (MG), oligodendrocytes (OD), and oligodendrocyte progenitor cells (OPC) as major cell types. Furthermore, microglia subtypes were further refined by projecting reference annotations from Green et al.,^19^ aligning their classification with our identified clusters. Consequently, the four major microglial subtypes, MG1–4, were defined as follows: MG1 (Mic. 2–5), containing surveilling genes (*CXCR1*⁺, *P2RY12*⁺); MG2 (Mic. 6–8), containing reactive genes (*TMEM163*⁺, *IL4R*⁺); MG3 (Mic. 9–10), enriched for redox-related genes; and MG4 (Mic. 12–13), characterized by lipid-processing genes (*APOE*⁺, *ADAM10*⁺, *GPNMB*⁺). Additional subtypes (MG5–8) were also identified but contained fewer cells and were not further analyzed in this study.

### Detection and characterization of cells with mCAs from brain snRNA-seq

We applied Numbat (v14.1)^18^ to the BAM files generated using 10x Genomics Cell Ranger,^28^ using cells that passed the above quality control, to detect cells carrying the same mCAs identified in blood WGS. Numbat integrated haplotype information with allele and expression signals from the single-cell sequencing data to detect copy number variants. The expression references in the brain were generated from each of the ROS/MAP snRNA-seq datasets. We specified the location of mCA segments from the blood WGS as the existing CNV profiles information, and ran Numbat with default parameters. Given the difficulty in detecting CNVs and the technical noise inherent in snRNA-seq data, we reasoned that enrichment of cells carrying the same CNVs as mCAs identified in blood among microglia-annotated cells would support the reliability of detecting blood-derived mCA-positive cells, assuming that microglia-annotated cells are the only ones potentially originating from blood. Accordingly, we considered mCAs to be reliably detected when the odds ratio for enrichment in microglia-annotated cells exceeded 3 and the *P*-value from Fisher’s exact test, Bonferroni-corrected by the number of mCA segments tested in each dataset, smaller than 0.05 was statistically significant.

To identify which previously defined microglial states^29^ are enriched among microglia-annotated cells harboring mCAs, we fit separate logistic regression models for each state. In each model, the presence of a given microglial state was treated as a binary variable of mCA status. A random effect for the sample was included to account for within-sample correlation, and log-transformed total UMI counts, disease status (i.e., AD or not), and study cohort were included as covariates. Only microglial states represented by more than five cells (i.e., MG1–4) were tested. *P*-values were Bonferroni-corrected based on the number of microglial states tested.

