## Supplementary Information for "Mosaic chromosomal alterations in blood are associated with an increased risk of Alzheimer’s disease"

### Table of Contents

#### Supplementary Figure

1. Genomic distribution of autosomal mCAs
2. Association of autosomal mCAs with AD stratified by *APOE* ε4 status in each ancestry group
3. Association of autosomal mCAs with AD conditioned on CHIP
4. Detection of the same mCA as identified in blood from brain snRNA-seq data in the replication dataset

### Supplementary Figure 1  Genomic distribution of autosomal mCAs

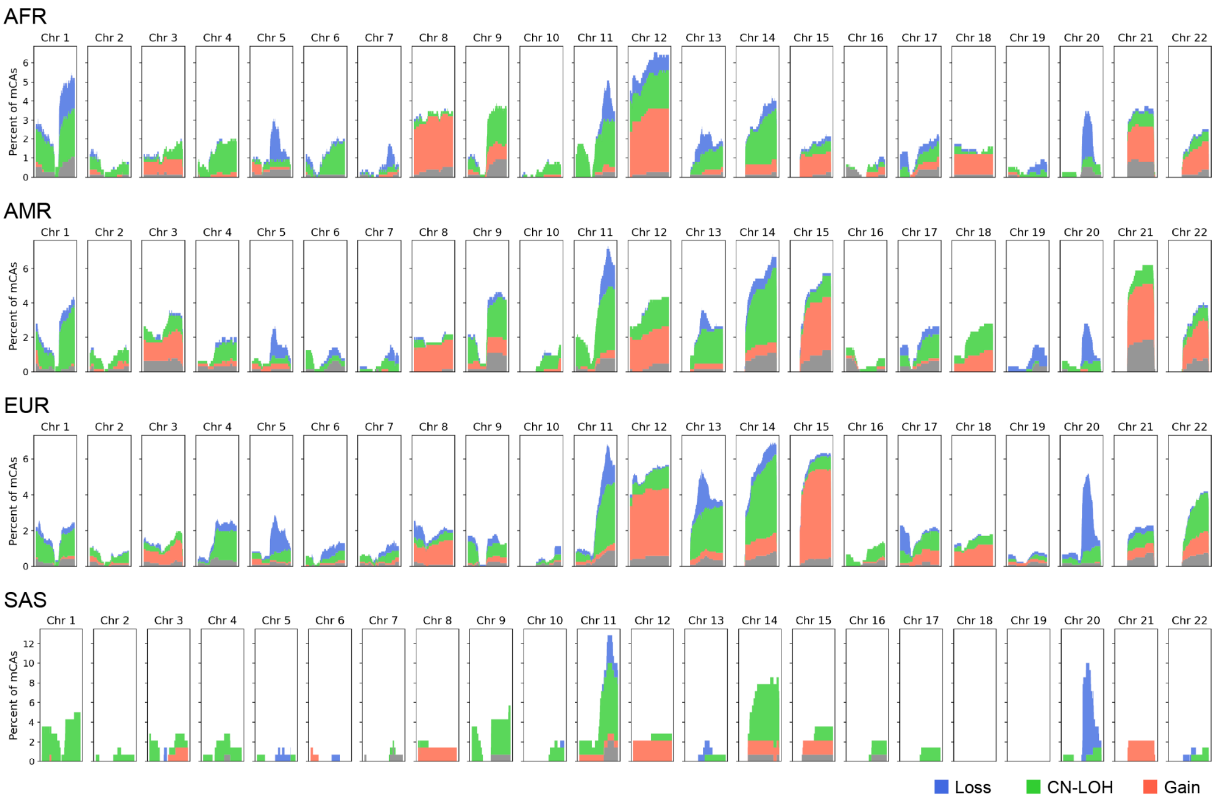


Histograms of mCA calls across the genome by chromosome for each genetic ancestry group. The horizontal axis represents 1 Kbp windows along each chromosome, and the vertical axis indicates the percentage of mCA calls spanning each genomic window. ALL, all ancestries; AFR, African American; AMR, Admixed American; EUR, European; SAS, South Asian; AD, Alzheimer's disease; OR, odds ratio.

**Supplementary Figure 2 Association of autosomal mCAs with AD stratified by APOE ε4 status in each ancestry group**

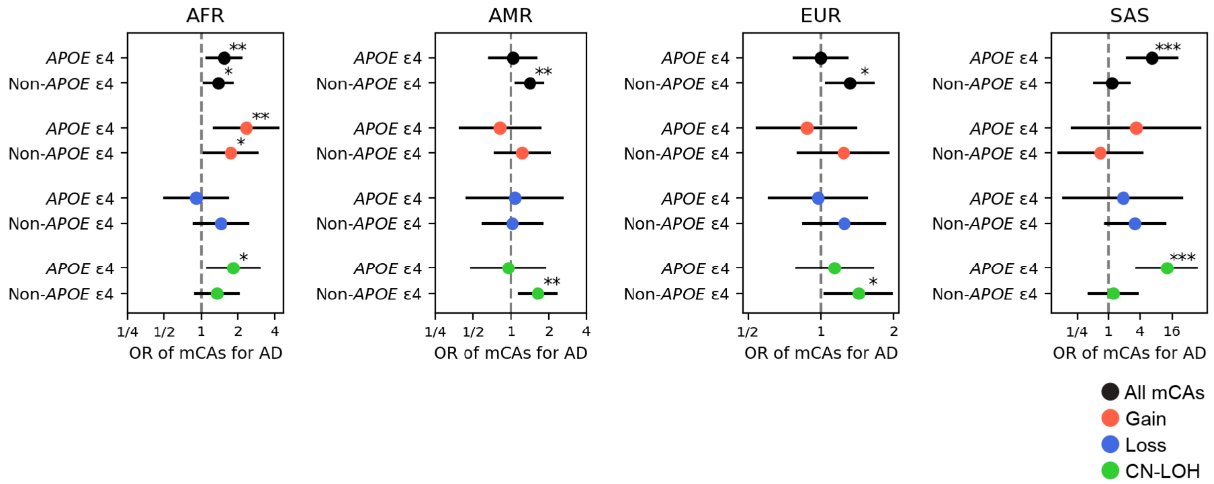


Forest plots showing the ORs of mCAs for AD in individuals with and without the APOE ε4 allele in each ancestry group.

In all plots, the error bars represent 95% confidence intervals. *, **, and *** represent *P*-value in association test < 0.05, 0.01, and 0.001, respectively.

ALL, all ancestries; AFR, African American; AMR, Admixed American; EUR, European; SAS, South Asian; AD, Alzheimer's disease; OR, odds ratio.

**Supplementary Figure 3  Association of autosomal mCAs with AD conditioned on CHIP**

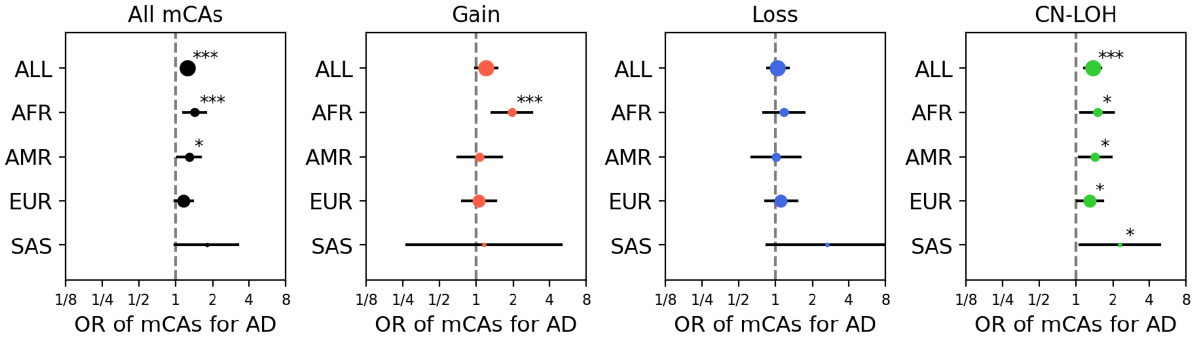


Forest plots showing the odds ratio (ORs) of mCAs for AD in each ancestry group conditioned on the presence of CHIP, with results presented by mCA type.

In all plots, the error bars represent 95% confidence intervals. *, **, and *** represent *P*-value in association test < 0.05, 0.01, and 0.001, respectively.

ALL, all ancestries; AFR, African American; AMR, Admixed American; EUR, European; SAS, South Asian; AD, Alzheimer's disease; OR, odds ratio.

**Supplementary Figure 4 Detection of the same mCA as identified in blood from brain snRNA-seq data in the replication dataset**

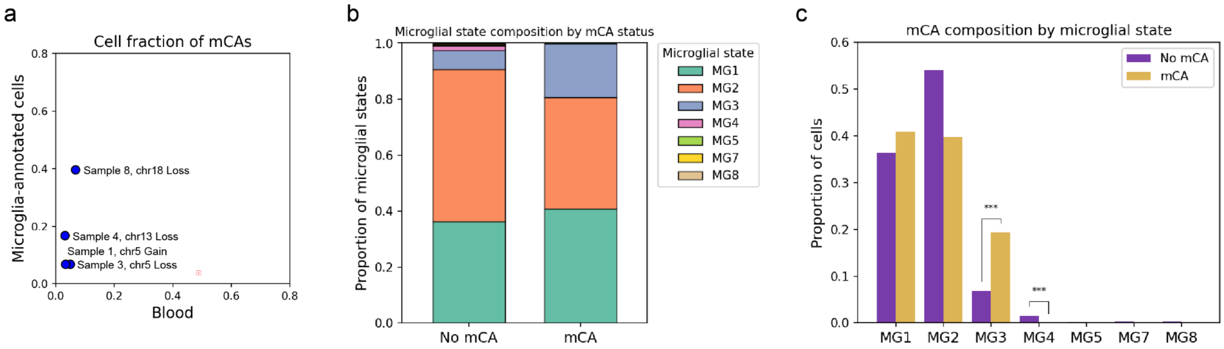


**a.** Scatter plot showing the cell fractions of mCAs in blood and microglia-annotated cells. **b.** Bar plot showing the proportion of microglial states by mCA status. **c.** Bar plot showing the proportion of cells with and without mCA across different microglial states.
